# Lipoprotein(a) and cardiovascular disease: prediction, attributable risk fraction and estimating benefits from novel interventions

**DOI:** 10.1101/2020.03.30.20043554

**Authors:** Paul Welsh, Claire Welsh, Carlos A Celis-Morales, Rosemary Brown, Lyn D Ferguson, Patrick B Mark, James Lewsey, Stuart R Gray, Donald M Lyall, Jason MR Gill, Jill P Pell, James A de Lemos, Peter Willeit, Naveed Sattar

## Abstract

**Background:** Lipoprotein (a) (Lp(a)) is a CVD risk factor amenable to intervention and might help guide risk prediction.

**Objectives:** To investigate the population attributable fraction due to elevated Lp(a) and its utility in risk prediction.

**Methods:** Using a prospective cohort study, 413,724 participants from UK Biobank, associations of serum Lp(a) with composite fatal/nonfatal CVD (n=10,065 events), fatal CVD (n=3247), coronary heart disease (n=16,649), ischaemic stroke (n=3191), and peripheral vascular disease (n=2716) were compared using Cox models. Predictive utility was determined by C-index changes. The population attributable fraction was estimated.

**Results:** Median Lp(a) was 19.7nmol/L (interquartile interval 7.6-75.3nmol/L). 20.8% had Lp(a) values >100nmol/L; 9.2% had values >175nmol/L. After adjustment for classical risk factors, in participants with no baseline CVD and not taking a statin, 1 standard deviation increment in log Lp(a) was associated with a HR for fatal/nonfatal CVD of 1.09 (95%CI 1.07-1.11). Associations were similar for fatal CVD, coronary heart disease, and peripheral vascular disease. Adding Lp(a) to a prediction model containing traditional CVD risk factors improved the C-index by +0.0017 (95% CI 0.0009, 0.0026). We estimated that having Lp(a) values >100nmol/L accounts for 5.7% of CVD events in the whole cohort. We modelled that an ongoing trial to lower Lp(a) in patients with CVD and Lp(a) above ∼175nmol/L may reduce CVD risk by 20.3%, assuming causality, and an achieved Lp(a) reduction of 80%.

**Conclusions:** Population screening for elevated Lp(a) may help to predict CVD and target Lp(a) lowering drugs, if such drugs prove efficacious, to those with markedly elevated levels.

## Introduction

Lipoprotein (a) (Lp(a)) is a low-density lipoprotein particle made by the liver, comprised of both an apolipoprotein(a) and an apolipoprotein B protein. Its structure is highly heterogeneous, but levels of Lp(a) are 80%-90% genetically determined and relatively stable across the life-course.

Epidemiological evidence shows strong associations of circulating Lp(a) with atherogenesis and consequent risk of cardiovascular disease (CVD). For instance, in a recent meta-analysis of statin trial data, those with Lp(a) concentrations above 50mg/dl were at 35% higher risk of incident CVD events (95%CI 11-66%) compared to those with Lp(a) <15mg/dl (approx. 30nmol/L) after adjusting for confounders (1). Similar data have been reported in population studies (2, 3). Furthermore, genetic data as well as basic science support the notion that the association is causal (4). This has led to interest both in the potential for Lp(a) to serve as a biomarker to enhance CVD risk prediction (5), and as a therapeutic target. Indeed, Lp(a) lowering drugs may be a viable therapeutic option, with at least one drug moving to phase 3 in the Lp(a) HORIZON trial (6). Further, proprotein convertase subtilisin/kexin type 9 (PCSK9) inhibitors, which lower Lp(a) concentrations by around 27%, may be particularly beneficial in reducing risk in patients with previous CVD and substantially raised Lp(a) (7).

Currently, most guidelines and consensus statements do not advocate widespread screening for elevated Lp(a) and suggest focus should be on measuring Lp(a) in people at high risk of CVD or in those in whom risk is intermediate when high Lp(a) would serve as a risk enhancer (8–10). However, the recently released ESC/EAS guidelines suggested a “one-off measurement of Lp(a) may help to identify people with very high inherited Lp(a) levels who may have a substantial lifetime risk of atherosclerotic cardiovascular disease (ASCVD)” (11). Although Lp(a) is sometimes measured in patients with suspected familial hypercholesterolaemia (12), it is currently not routinely measured in general practice (9). In addition, there is conflicting advice on what constitutes a “high” Lp(a) level. Several guidelines and consensus statements advocate the 50mg/dl (∼125nmol/L) cutoff (8, 9, 13) as this corresponds to the 80^th^ percentile in one cohort study (14), but the 2016 Canadian Cardiovascular Society Guidelines use 30mg/dl on the basis of elevated CVD risk (∼75nmol/L) (15). The Lp(a) HORIZON trial uses 70mg/dl (∼175nmol/L) as an inclusion criterion (16). The lack of data from a single large cohort with consistent phenotyping is a significant limitation in interpreting the existing literature, impacting our understanding of the prevalence of high Lp(a), and its consequences for CVD risk.

UK Biobank is a large prospective population-based cohort study carried out in the UK, with information on baseline biochemistry measurements including routine lipids and Lp(a) measured in core laboratories. We aimed to use this resource to explore the shape of the association of Lp(a) with a range of distinct CVD outcomes to investigate the population attributable risk fraction for CVD that might be explained by elevated Lp(a), and to predict what might be the effect of novel Lp(a)-lowering therapies based on these data and recent relevant trials.

## Methods

UK Biobank was conducted across 22 assessment centres across the UK between April 2007 and December 2010 and recruited 502,624 participants aged 37 to 73. Baseline biological measurements were recorded and touch-screen questionnaires were administered according to a standardised protocol (17, 18). UK Biobank received ethical approval from the North West Multi-Centre Research Ethics Committee (REC reference: 11/NW/03820). All participants gave written informed consent before enrolment in the study, which was conducted in accord with the principles of the Declaration of Helsinki.

For the present analysis, ethnicity was coded as white, South Asian, black, or mixed/other. Smoking status was categorised into never or former/current smoking. Systolic and diastolic blood pressure were measured at the baseline visit, preferentially using an automated measurement, but using manual measurement where this was not available. Blood collection sampling procedures for the study have been previously described and validated (19). The definition of baseline diabetes included self-reported type 1 or type 2 diabetes, those with a primary or secondary hospital diagnoses relating to diabetes at baseline (ICD-10 codes E10-E14.9), and those who reported using diabetes medications. Statin (categorised to include other cholesterol lowering medications) and blood pressure medication use were also recorded from self-report. Baseline cardiovascular disease was defined as self-reported myocardial infarction, stroke, or transient ischaemic attack.

Biochemistry measures were performed at a dedicated central laboratory on around 480,000 samples between 2014 and 2017. These included serum total cholesterol and HDL cholesterol (Beckman Coulter, UK on an AU5800 platform) and Lp(a) (Randox Bioscience, UK on an AU5800 platform). Data were adjusted by UK Biobank centrally before release to adjust for pre-analytical variables. For Lp(a), low, medium, and high-quality control materials ran with coefficients of variation of ≤6.1%. Further details of these measurements can be found in the UK Biobank online showcase and protocol (http://www.ukbiobank.ac.uk). Lp(a) was reported in nmol/L rather than mg/dl, reflecting the concentration of particles rather than the mass of particles. The minimum reported concentration of Lp(a) was 3.8nmol/L and the maximum was 189nmol/L; participants who had levels below the lower level (n=48,360) were coded as having an Lp(a) concentration of 2.88nmol/L, and above the upper level (n=34,195) coded as 250nmol/L for continuous analyses.

Date and cause of death were obtained from death certificates held by the National Health Service (NHS) Information Centre for participants from England and Wales and the NHS Central Register Scotland for participants from Scotland. Only primary causes of death listed on the death certificate were included in this analysis. Nonfatal outcomes were ascertained by linkage of participant study data to Hospital Episode Statistics (HES) from the National Health Service. The primary outcome of interest was ASCVD, a composite CVD outcome that reflects the ACC/AHA guidelines prediction score including death from CHD or stroke (ICD-10 I20-25, I60-64) or hospitalisation for myocardial infarction or stroke (ICD-10 I21, I22, I60, I61, I62, I64) (20). A secondary outcome was fatal cardiovascular disease as defined by primary cause of death from events included in the European SCORE clinical guidelines (I10-15, I44-51, I20-25, I61-73) (21). We also investigated secondary outcomes of fatal or nonfatal coronary heart disease (ICD10 I20-25, or operation code K40-49, K50, K75, U19)), peripheral vascular disease (ICD10 I70-78, or operation code K551), heart failure (ICD10 I150, I42.0, I42.6, I42.7, I42.9, I11.0), and ischaemic stroke (ICD10 I63, I64).

End of follow-up for each participant was recorded as the date of death or the date of end of follow-up for the assessment centre attended, whichever came first. The period at risk of each participant began on the date of their assessment.

### Statistical analyses

The association of continuous log-transformed Lp(a) with other lipid variables was tested using Pearson correlation coefficients. Lp(a) was analysed using a number of different models, reflecting existing uncertainty regarding cut-offs for “abnormal” levels. Lp(a) was categorised into groups with cutoffs at 20nmol/L and 100nmol/L or 125nmol/or 150nmol/L or 175nol/L. Sex and ethnicity specific centiles (50^th^, 75^th^, 80^th^, 90^th^ and 95^th^ centiles) for Lp(a) were also developed, using binomial exact confidence intervals to yields 95% confidence intervals (CI) for the centiles. The sex and ethnicity specific 80^th^ centile was chosen to create a binary “high” category for Lp(a). Log transformed Lp(a) was also analysed as a continuous variable. Classical risk factors were expressed as mean (standard deviation) if symmetrically distributed, median (interquartile range) if skewed, and number (%) if categorical. The distribution of classical risk factors by categories of outcome or exposure of interest were assessed using unpaired 2 tail t-tests, a Wilcoxon rank-sum, or a chi-squared test, respectively.

The cohort was analysed as a whole cohort, and was also stratified as a primary prevention cohort (participants without baseline CVD and not taking a statin) and as a high risk cohort (participants with baseline CVD and/or taking a statin). Rates of the primary composite CVD outcome were investigated in unadjusted models, splitting the cohort by the specified Lp(a) categories. Associations of continuous and categorical Lp(a) with outcomes of interest were investigated using Cox-proportional hazard models in the whole cohort adjusting for classical risk factors (age, sex, total cholesterol, HDL cholesterol, ethnicity, smoking, systolic blood pressure, blood pressure medications, baseline diabetes), and also adjusting for baseline CVD and baseline statin use in analyses including high risk participants. The proportional hazard assumption was checked by inspection of Schöenfeld residuals. We checked for interactions of Lp(a) effect with median age, sex, ethnicity, baseline CVD, statin use, and high total cholesterol (at 8.0mmol/L). Associations of Lp(a) with outcomes of interest were then conducted separately in the primary prevention and high risk cohorts. We explored the shape of the association of Lp(a) with outcomes using restricted cubic splines, and using linear and categorical models. Follow-up time was calculated as days from enrolment to death or the end of follow-up, whichever occurred first. Analyses were repeated in the group of people with baseline CVD or statin use at baseline, additionally adjusting for baseline CVD and statin use.

The ability of Lp(a) to improve prediction of CVD was tested by assessing improvement over a base model containing all elements from the Pooled Cohort Equation. The model for the composite CVD outcome tested whether Lp(a) improved prediction after inclusion of classical risk factors of age, sex, ethnicity, total cholesterol, HDL cholesterol, systolic blood pressure, diabetes, smoking, statin and blood pressure medication use. Improvement in prediction was tested using Harrells C-Index for survival data, testing for increased concordance after the addition of Lp(a) to the model as either a continuous or categorical variable. We used a categorical net reclassification index (NRI), to investigate the changes in predicted risk classification after addition of Lp(a) to models, using a pre-specified binary 10-year CVD risk boundary of 7.5%, reflecting high or low risk categorisation according to guidelines.

Population attributable fractions in the exposed, with 95% confidence intervals, were estimated using two adjusted Cox models and the punafcc postestimate command in STATA. The first model added Lp(a) as a binary variable (detectable/undetectable), and was designed to estimate the causal effect of Lp(a) in the whole cohort. The second model added Lp(a) as a 4-category variable with concentration >175nmol/L (representing the Lp(a) HORIZON trial recruitment criterion (6)), 40-175nmol/L, 30-40nmol/L (the estimated attained Lp(a) assuming an 80% reduction on-treatment (22)), and <30nmol/L. This model was then used to test the estimated proportional reduction in CVD events among those with Lp(a) >175nmol/L when concentration was lowered to the 30-40nmol/L range. The model was run in the whole cohort and in those with baseline CVD.

All analyses were performed using STATA 14 (StataCorp LP) and R (version 3.5.1).

## Results

### Cross sectional associations

Of 502,624 people included in the study, complete data on covariates, including Lp(a) were available in 413,724 participants. Median Lp(a) in the cohort was 19.7nmol/L (interquartile interval 7.6-75.3nmol/L) and in participants without baseline CVD and not taking a statin this was 19.1nmol/L (interquartile interval 7.6-70.5nmol/L). The 80^th^ centile in the whole cohort was 104.5nmol/L (95% CI 103.8-105.3). In the whole cohort, 85,930 (20.8%) had Lp(a) above 100nmol/L, 68,601 (16.6%) above 125nmol/L, 52,157 (12.6%) above 150nmol/L, and 38,109 (9.2%) above 175nmol/L. Sex and ethnicity specific cut-offs show that women and participants with black ethnicity more frequently had elevated Lp(a) (Figure 1).

**Figure 1.**
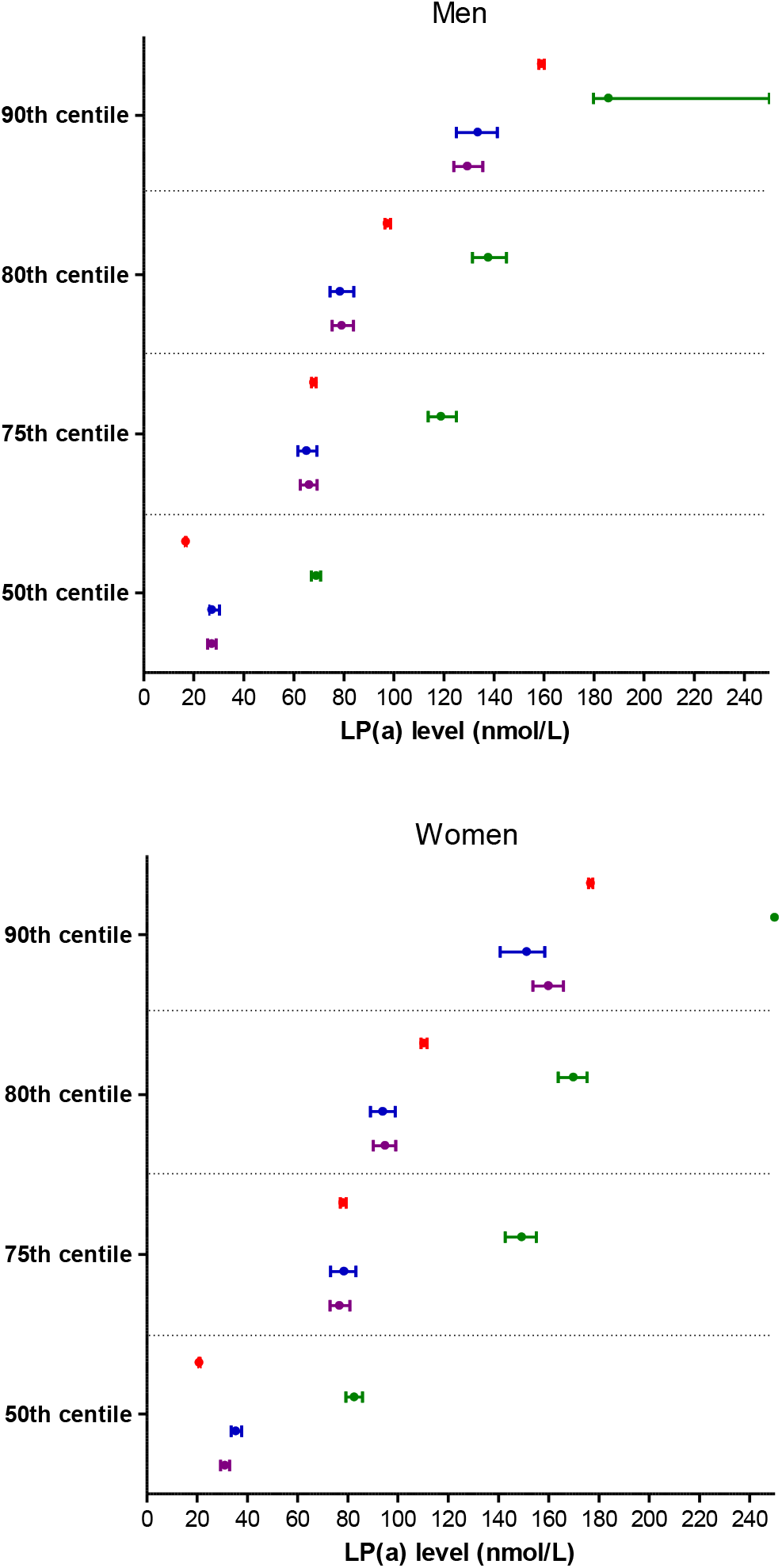
Centiles of Lp(a), along with 95% confidence intervals, by sex and ethnicity in the whole cohort. Red denotes white ethnicity, green denotes black ethnicity, blue denotes South Asian, and purple denotes other or mixed ethnicity.

Lp(a) had weak positive associations with total cholesterol (r=0.11), HDL-cholesterol (r=0.04), and LDL cholesterol (r=0.12) (p for all<0.0001). These associations were only nominally stronger in the population who did not report taking statins (r=0.14, 0.05, 0.16, respectively). Participants with elevated Lp(a) were generally slightly older, had slightly higher systolic blood pressure and total cholesterol, and were more likely to have baseline CVD (Table 1).

**Table 1.**
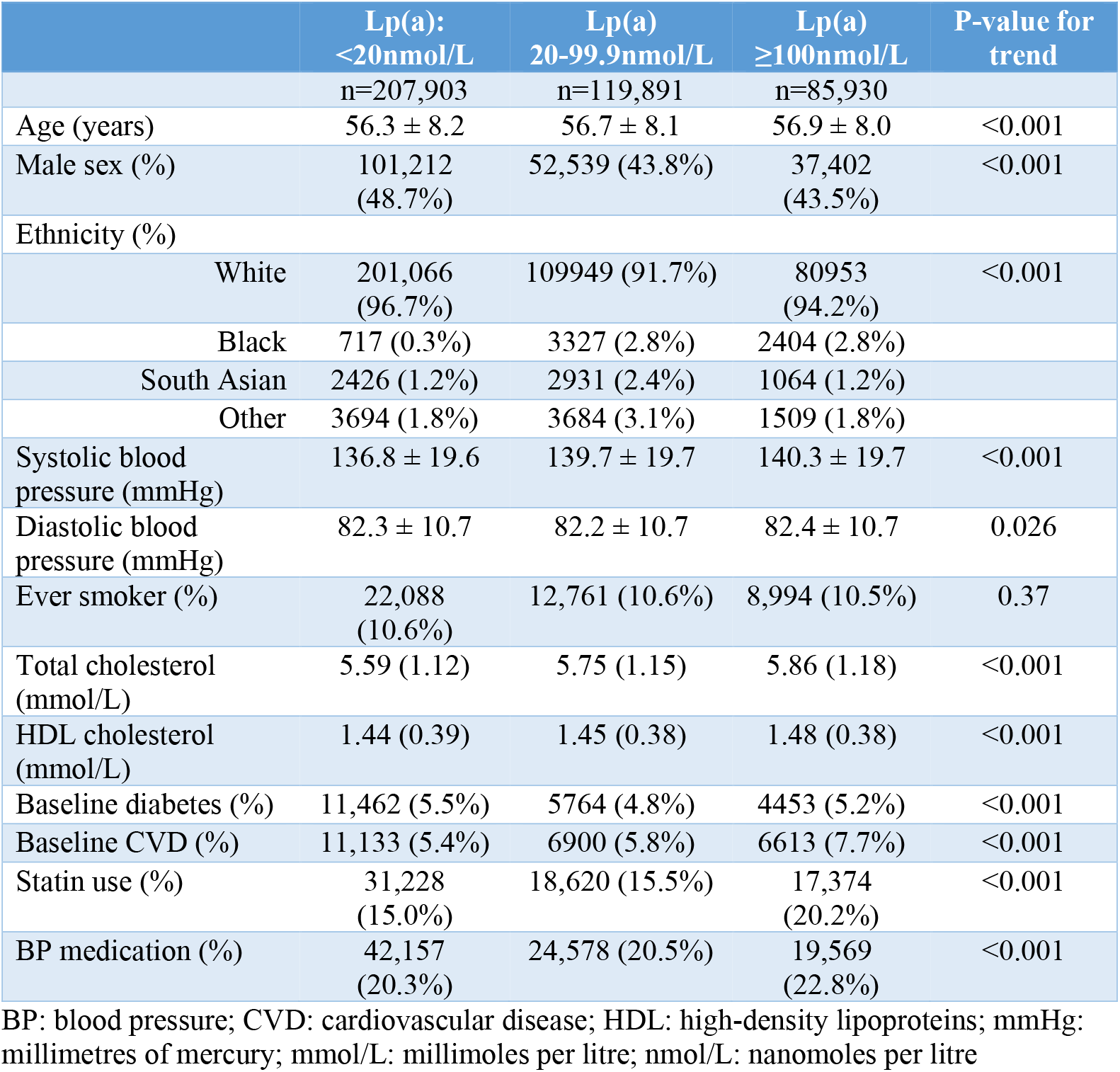
Association of categories of Lp(a) with classical risk factors for CVD at baseline (n=413,724)

|  | Lp(a):<br><20nmol/L<br>n=207,903 | Lp(a)<br>20-99.9nmol/L<br>n=119,891 | Lp(a)<br>≥100nmol/L<br>n=85,930 | P-value for<br>trend |
| --- | --- | --- | --- | --- |
| Age (years) | 56.3 ± 8.2 | 56.7 ± 8.1 | 56.9 ± 8.0 | <0.001 |
| Male sex (%) | 101,212<br>(48.7%) | 52,539 (43.8%) | 37,402<br>(43.5%) | <0.001 |
| Ethnicity (%) |  |  |  |  |
| White | 201,066<br>(96.7%) | 109949 (91.7%) | 80953<br>(94.2%) | <0.001 |
| Black | 717 (0.3%) | 3327 (2.8%) | 2404 (2.8%) |  |
| South Asian | 2426 (1.2%) | 2931 (2.4%) | 1064 (1.2%) |  |
| Other | 3694 (1.8%) | 3684 (3.1%) | 1509 (1.8%) |  |
| Systolic blood pressure (mmHg) | 136.8 ± 19.6 | 139.7 ± 19.7 | 140.3 ± 19.7 | <0.001 |
| Diastolic blood pressure (mmHg) | 82.3 ± 10.7 | 82.2 ± 10.7 | 82.4 ± 10.7 | 0.026 |
| Ever smoker (%) | 22,088<br>(10.6%) | 12,761 (10.6%) | 8,994 (10.5%) | 0.37 |
| Total cholesterol (mmol/L) | 5.59 (1.12) | 5.75 (1.15) | 5.86 (1.18) | <0.001 |
| HDL cholesterol (mmol/L) | 1.44 (0.39) | 1.45 (0.38) | 1.48 (0.38) | <0.001 |
| Baseline diabetes (%) | 11,462 (5.5%) | 5764 (4.8%) | 4453 (5.2%) | <0.001 |
| Baseline CVD (%) | 11,133 (5.4%) | 6900 (5.8%) | 6613 (7.7%) | <0.001 |
| Statin use (%) | 31,228<br>(15.0%) | 18,620 (15.5%) | 17,374<br>(20.2%) | <0.001 |
| BP medication (%) | 42,157<br>(20.3%) | 24,578 (20.5%) | 19,569<br>(22.8%) | <0.001 |

BP: blood pressure; CVD: cardiovascular disease; HDL: high-density lipoproteins; mmHg: millimetres of mercury; mmol/L: millimoles per litre; nmol/L: nanomoles per litre

### Univariable association of Lp(a) with outcomes

Median follow-up time for the composite CVD outcome was 8.9 years (IQR 8.2-9.5) in both the whole cohort and among participants in the cohort without baseline CVD and not taking a statin. The composite CVD outcome occurred in 10,065 participants (2.4%), and fatal CVD occurred in 3247 participants (0.8%) in the whole cohort. The composite CVD outcome occurred in 6125 participants (1.8%), and fatal CVD occurred in 1627 participants (0.5%) in the subgroup without baseline CVD and not taking a statin.

Baseline Lp(a) was higher among the participants who went on to experience the composite CVD outcome and the fatal CVD outcome (Supplementary Table 1). Lp(a) was also higher among those who went on to experience CHD or peripheral vascular disease (PVD), but was not higher among those who went on to experience ischaemic stroke, heart failure (Supplementary Table 1). Rates of the primary composite CVD outcome were higher in participants with higher categories of baseline Lp(a) (Figure 1).

### Multivariable association of Lp(a) with outcomes

Among those without baseline CVD and not taking a statin, after adjusting for classical risk factors, the shape of the association of Lp(a) with composite CVD, fatal CVD, and CHD was positive and broadly linear (Figure 2). There was also a weaker positive association with PVD, but no evidence of an association with stroke, heart failure or atrial fibrillation (Figure 3)

**Figure 2.**
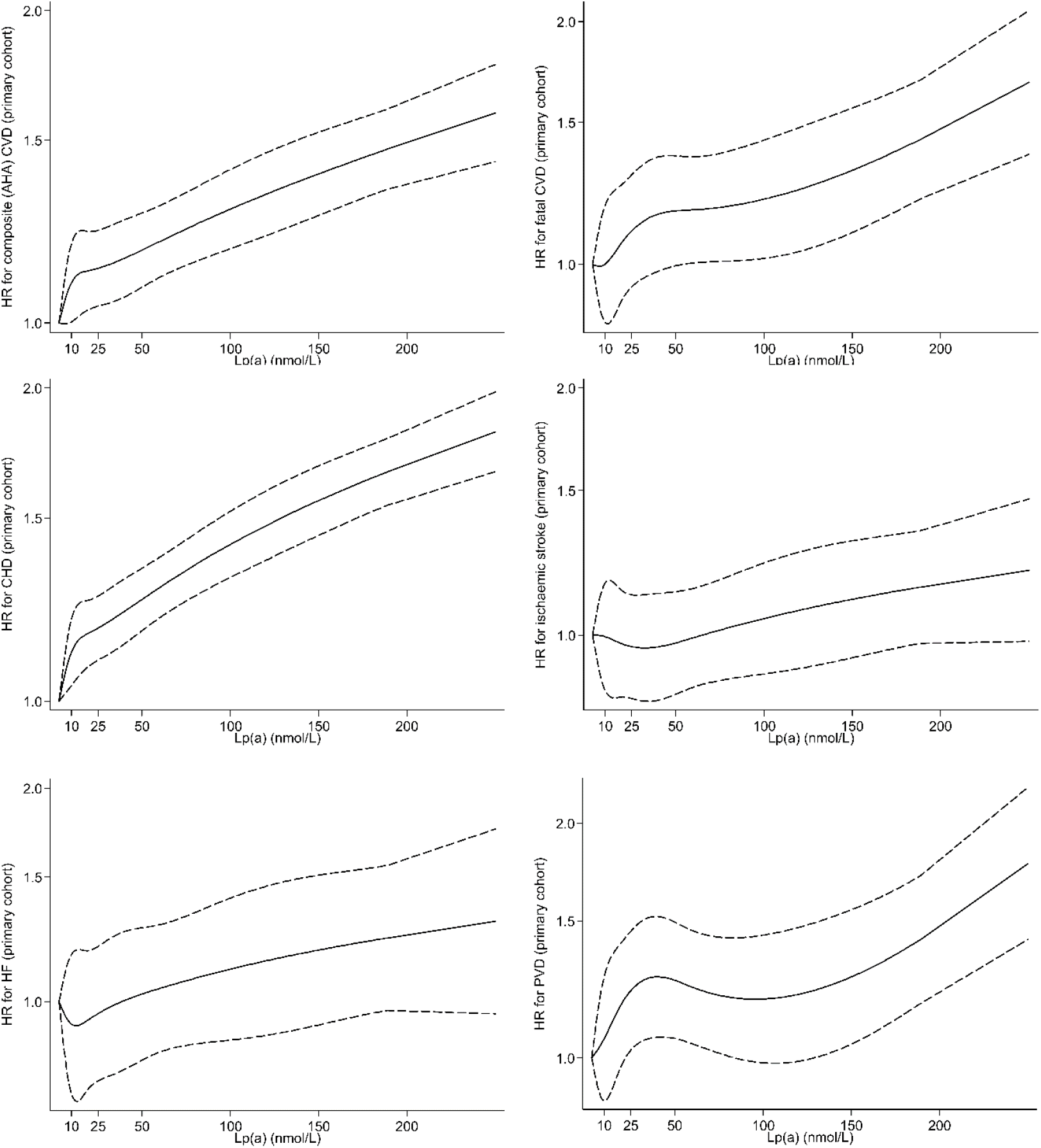
Association of Lp(a) with outcomes after adjusting for classical risk factors among participants without baseline CVD and not taking a statin.

**Figure 3.**
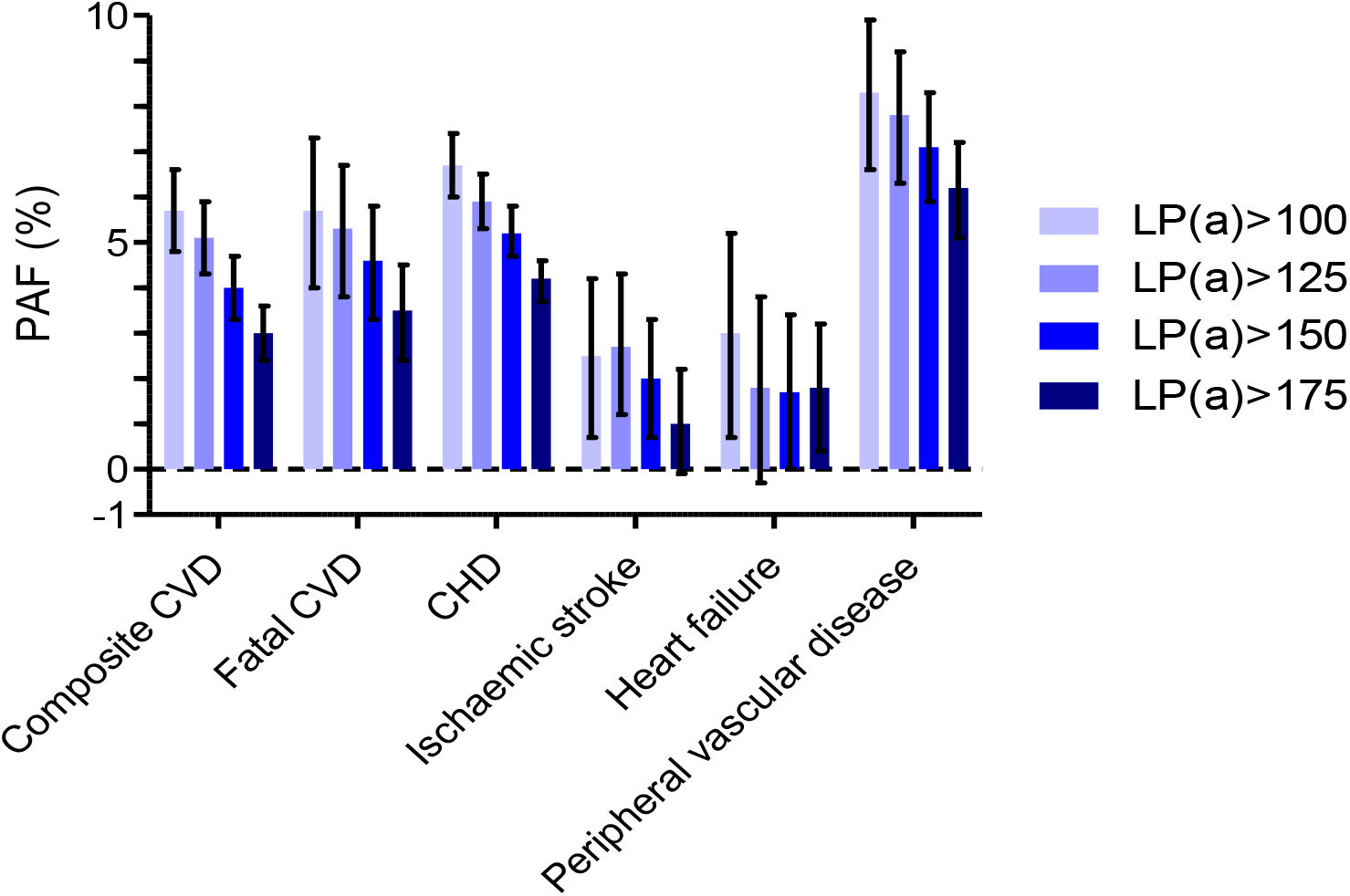
Population attributable fractions (with 95% confidence intervals) of Lp(a) over a range of thresholds for each outcome of interest within the whole cohort.

In the whole cohort, there was an independent association of one standard deviation increase in log Lp(a) with the primary composite CVD outcome (HR 1.09 (95% CI 1.07-1.11) after adjusting for classical risk factors statin use and baseline CVD. There was no interaction with age (above or below the median of 57 years) (p=0.11), sex (p=0.40), ethnicity (p>0.18 for each ethnic group compared to white), baseline CVD (p=0.12), statin use (p=0.23), or total cholesterol (above or below 8.0mmol/L cut-off) (p=0.34).

Although there was no formal interaction, for clarity data were split into the primary prevention cohort and the high risk cohort to further explore associations with outcomes. Using continuous and categorical models of Lp(a) there was a positive association with composite primary CVD outcome (Table 2). Lp(a) was also associated with fatal CVD, CHD and PVD as well as demonstrating a borderline association with ischaemic stroke after adjusting for classical risk factors, in both the primary prevention cohort and the high risk cohort (Supplementary Table 2). There was no association with heart failure (Supplementary Table 2).

**Table 2.** Association of Lp(a) (hazard ratio and 95% CI) as a continuous variable (per 1 SD increase in log Lp(a)) and as a categorical variable with the primary composite CVD outcome after adjusting for classical risk factors*.

| Lp(a) conc. | Primary prevention cohort |  | High risk cohort** |  |
| --- | --- | --- | --- | --- |
|  | N<br>(n events) | HR (95% CI) | N<br>(n events) | HR (95% CI) |
| Per 1SD | 340333<br>(6125) | 1.09<br>(1.07-1.11) | 73391 (3940) | 1.07<br>(1.05-1.09) |
| <20nmol/L | 173636<br>(2857) | Ref | 34267<br>(1688) | Ref |
| 20-99.9nmol/L | 99496<br>(1802) | 1.11<br>(1.04-1.18) | 20395<br>(1086) | 1.09<br>(1.01-1.17) |
| ≥100nmol/L | 67201<br>(1466) | 1.36<br>(1.28-1.45) | 18729<br>(1166) | 1.28<br>(1.18-1.37) |
| <20nmol/L | 173636<br>(2857) | Ref | 34267<br>(1688) | Ref |
| 20-124.9nmol/L | 114090<br>(2086) | 1.12<br>(1.06-1.19) | 23130<br>(1241) | 1.09<br>(1.01-1.18) |
| ≥125nmol/L | 52607<br>(1182) | 1.40<br>(1.31-1.50) | 15994<br>(1011) | 1.31<br>(1.21-1.41) |
| <20nmol/L | 173636<br>(2857) | Ref | 34267<br>(1688) | Ref |
| 20-149.9nmol/L | 127633<br>(2371) | 1.14<br>(1.08-1.21) | 26031<br>(1428) | 1.11<br>(1.03-1.19) |
| ≥150nmol/L | 39064<br>(897) | 1.43<br>(1.33-1.55) | 13093<br>(824) | 1.32<br>(1.21-1.43) |
| <20nmol/L | 173636<br>(2857) | Ref | 34267<br>(1688) | Ref |
| 20-174.9nmol/L | 138833<br>(2626) | 1.16<br>(1.10-1.23) | 28879<br>(1608) | 1.13<br>(1.05-1.21) |
| ≥175nmol/L | 27864<br>(642) | 1.44<br>(1.32-1.58) | 10245<br>(644) | 1.33<br>(1.21-1.45) |
| Below the sex and ethnicity specific 80<sup>th</sup> centile | 275886<br>(4672) | Ref | 54884<br>(2767) | Ref |
| Above the sex and ethnicity specific 80<sup>th</sup> centile | 64447<br>(1453) | 1.31<br>(1.24-1.39) | 18507<br>(1173) | 1.24<br>(1.16-1.33) |

\*age, sex, total cholesterol, HDL cholesterol, ethnicity, smoking, systolic blood pressure, blood pressure medications, baseline diabetes
\*\* Participants with baseline CVD or taking a statin; additionally adjusted for baseline CVD and statin use
CI: confidence interval; CVD: cardiovascular disease; HDL: high-density lipoproteins; HR: hazard ratio; Lp(a): Lipoprotein (a); nmol/L: nanomoles per litre; SD: standard deviation

### Lp(a) and prediction of CVD

Prediction of incident CVD was specifically explored in the primary prevention cohort. In a model of CVD prediction based on pooled cohort equation risk factors plus current treatments, classical risk factors yielded a C-index of 0.7439 (95% CI 0.7381-0.7497). Addition of Lp(a) as a continuous variable to these risk factors increased the C-index by +0.0017 (95%CI 0.0009, 0.0026) (Table 3). On addition of Lp(a) to the model the improvement in the categorical NRI was 1.39% (95% CI 0.69-2.12%) and most of the improvement was due to up classification of risk among cases (Table 3). Similar small improvements in prediction were obtained when Lp(a) was added as a categorical variable, with no clear advantage of one model over another (Table 3).

**Table 3.** Improvement in prediction of CVD among participants in the primary prevention cohort measured by the C-statistic and categorical net reclassification index (across the 7.5% 10-year risk boundary).

| Model | C-Index | Change in C-Index | Overall NRI | Case NRI | Control NRI |
| --- | --- | --- | --- | --- | --- |
| Classical risk factors* | 0.7439<br>(0.7381, 0.7497) |  |  |  |  |
| Classical risk factors + continuous log Lp(a) |  | +0.0017<br>(0.0009, 0.0026) | +1.39%<br>(0.69, 2.12%) | +1.46%<br>(0.77, 2.19%) | -0.08%<br>(-0.13, -0.03%) |
| Classical risk factors + categorical Lp(a) at <20, 20-99.9, and >100nmol/L |  | +0.0018<br>(0.0009, 0.0026) | +0.98%<br>(0.26, 1.71%) | +1.07%<br>(0.37, 1.79%) | -0.09%<br>(-0.14, -0.05%) |
| Classical risk factors + categorical Lp(a) at <20, 20-149.9, and >150nmol/L |  | +0.0016<br>(0.0007, 0.0024) | +1.15%<br>(0.42, 1.88%) | +1.23%<br>(0.51, 1.96%) | -0.08%<br>(-0.13, -0.03%) |
| Classical risk factors + binary Lp(a) at sex and ethnicity specific 80<sup>th</sup> percentile |  | +0.0015<br>(0.0007, 0.0023) | +1.15%<br>(0.52, 1.80%) | +1.21%<br>(0.57, 1.86%) | -0.06%<br>(-0.11, -0.01%) |

\*age, sex, total cholesterol, HDL cholesterol, ethnicity, smoking, systolic blood pressure, blood pressure medications, baseline diabetes
CVD: cardiovascular disease; HDL: high-density lipoproteins; Lp(a): Lipoprotein (a); nmol/L: nanomoles per litre; NRI: net reclassification index

### Population attributable fraction of Lp(a)

After adjusting for confounders, the proportion of CVD attributable due to any detectable Lp(a) in the whole cohort was 8.6% (95% CI 7.4-9.8%); this is the reduction in risk expected if the whole cohort had Lp(a) below the limit of detection.

In the whole cohort, an Lp(a) above 100nmol/L accounted for 5.7% (95% CI 4.8, 6.6%) of the composite CVD outcome (Figure 3). Lp(a) elevation above 100nmol/L accounted for 5.7% (4.0, 7.3%) of fatal CVD, 6.7% (6.0, 7.4%) of CHD, 2.5% (0.7, 4.2%) of stroke, 3.0% (0.7, 5.2%) of heart failure, and 8.3% (6.6, 9.9%) of peripheral vascular disease. Of these outcomes, the only one where the Lp(a) attributable fraction varied substantially in the high risk cohort compared to the primary prevention cohort was peripheral vascular disease, with a population attributable fraction of 4.7% (2.4, 7.0%) in the primary prevention cohort and 12.1% (9.6, 14.6%) in the high risk cohort. Moving the threshold for “high” Lp(a) to higher cutoffs resulted in somewhat lower, but still substantial, attributable fractions due to reduced prevalence of the higher cutoffs (Figure 3). The overall PAF lowered to 3.0% (95%CI 2.4-3.6%) for a cutoff of Lp(a) above 175nmol/L due to reduced prevalence.

### Expected benefit of Lp(a) reduction

We then specifically modelled the scenario in the ongoing phase 3 trial of an Lp(a) lowering agent. For all participants with an Lp(a) above 175nmol/L, reducing this by approximately 80% so that participants have Lp(a) 30-40nmol/L range, yields an estimated risk reduction of 23.2% (95%CI 15.0-30.6%). For participants with baseline CVD and an Lp(a) above 175nmol/L, similar Lp(a) reductions are estimated to reduce risk by 20.3% (95%CI 2.5-34.9%).

## Discussion

In this large cohort of over 400,000 individuals, a high proportion of the UK biobank cohort had what might conventionally be called high Lp(a) levels; 20.8% above 100nmol/L and 9.2% above 175nmol/L. We noted a broadly linear relationship of Lp(a) with composite fatal or nonfatal CVD, fatal CVD, and fatal or nonfatal CHD, with associations largely unaffected by other risk factors. Therefore, population attributable fractions for Lp(a) were notable. Further, we estimate that targeting Lp(a) lowering therapy in ongoing trials to those with Lp(a) concentrations above 175nmol/L would reduce CVD incidence by around 20% (regardless of baseline CVD status). Collectively, our results seem to justify the recent ESC/EAS guidelines (11) suggesting consideration for at least a one-time Lp(a) measurement in all people being screened for cardiovascular disease.

Genetic data as well as basic science support the notion that Lp(a) causes CVD (4). Although genetic data suggested large reductions in Lp(a) would be required to show clinical benefit, recent phase 2 trial data show that the drug AKCEA-APO(a)-LRx (also called TQJ230) reduces Lp(a) substantially, with 80-90% reductions in patients with established CVD and high Lp(a) levels, depending on dosing (22)(23). This antisense oligonucleotide inhibits the production of apolipoprotein(a), thereby reducing Lp(a). Phase 3 outcome trials are underway (6) and specifically target those with elevated Lp(a). If this or other drugs targeting Lp(a) prove successful in reducing CVD events, population screening of Lp(a) may be one way to target those who are most likely to benefit. Our data suggest that a drug that prevents CVD through Lp(a) lowering may also have benefits for individual outcome components of the CVD composite, and for PVD outcomes. Our models suggest potential benefit in both primary and secondary prevention. Although Lp(a) only adds moderate information to risk discrimination metrics, and is more expensive than traditional lipid measurements (24), the fact that the marker is i) causal, ii) largely orthogonal to other risk factors, iii) stable across lifecourse, iv) has a substantial population attributable fraction, and v) may help guide therapy allocation, enhances arguments that measurement of Lp(a) should become more common in the evaluation of CVD risk. Notably, the reported improvement in C-statistics with Lp(a) was around four times higher than previously reported for CRP (25).

The ethnicity-specific centiles we report confirm and extend observations in other cohorts, most noticeably higher Lp(a) in black people (26). However, we note no interaction of Lp(a) with demographics or other risk factors suggesting that Lp(a) is similarly associated with risk in different subgroups. We would expect a higher PAF for CVD outcomes in black ethnicities due to higher prevalence of the exposure, but low numbers of black participants restrict our ability to formally test this hypothesis in this cohort. The PAFs we report for Lp(a) may usefully be put in context of estimates of PAFs for other risk factors. The ARIC study reported PAFs at examination four (among white participants) of 21% for hypertension, 13% for diabetes, 10% for hypercholesterolaemia and 12% for smoking (27). Similarly, in the Emerging Risk Factors Collaboration, the PAF for diabetes in vascular death has been estimated at 11% (assuming a 10% diabetes prevalence (28)), In terms of CVD risk prediction, our data are also broadly in line with the Emerging Risk Factors Collaboration (ERFC), where data from 165,544 participants in 37 prospective studies showed Lp(a) improved the C-index by +0.0016 (95% CI, 0.0009-0.0023) (29), lending strong external validity to these new reported findings. It is also in agreement with data from other large cohort studies (30–33). Our data extend these findings in a large cohort with substantial power, using a single method of Lp(a) measurement, where we also estimated the PAF for CVD outcomes independently due to Lp(a) elevation.

### Study strengths and limitations

The strengths of our study include the large cohort size at an age relevant to CVD risk scoring, and biochemistry assays performed in a single dedicated central laboratory. We were also able to extensively adjust our models for classical risk factors and separately analysed participants already on statins as well as those with previous CVD. We were also able to look at other cardiac outcomes such as heart failure. Potential limitations include the relatively low average CVD risk of participants, although risk prediction models performed broadly in line with expectations. UK Biobank is not representative of the whole UK population (30), and while this is generally not a concern in investigating risk associations (34) it will have an impact on calculated population attributable fractions. The population attributable fractions we observe here cannot be taken as representative of the UK population as a whole. However, due to the under-representation of black people in UK Biobank, it may be that our estimates are conservative. In addition, the 80^th^ centile we report here of 105nmol/L corresponds broadly to previously reported 80^th^ centiles in 3000 men and 3000 women from the Copenhagen General Population Study (50mg/dl) (9). Finally, our estimates of expected therapy effects among the exposed (Lp(a) above 175nmol/L) should be robust to differences in representativeness, since we only consider those with high Lp(a) as exposed.

## Conclusion

In conclusion, in this, the largest single prospective study of Lp(a) levels, our findings add strong support for recent guideline recommended one-time measurement of Lp(a) in CV risk assessment to identify a large proportion with markedly elevated levels sufficient to contribute to atherothrombotic risk. Our work also provides support to the ongoing programmes to develop efficacious Lp(a) lowering drugs

## Data Availability

Researchers can apply to use the UK Biobank resource and access the data used.

https://bbams.ndph.ox.ac.uk/ams/

## Abbreviations

ACC: American College of Cardiology
AF: atrial fibrillation
AHA: American Heart Association
ASCVD: atherosclerotic cardiovascular disease
CHD: coronary heart disease
CI: confidence interval
CVD: cardiovascular disease
EAS: European Atherosclerosis Society
ERFC: Emerging Risk Factors Collaboration
ESC: European Society of Cardiology
HDL: high-density lipoproteins
HES: Hospital Episode Statistics
HF: heart failure
HR: hazard ratio
ICD: International Classification of Diseases
LDL: low-density lipoproteins
Lp(a): Lipoprotein (a)
mmol/L: millimoles per litre
NHS: National Health Service
nmol/L: nanomoles per litre
NRI: net reclassification index
PAF: paroxysmal atrial fibrillation
PCSK9: Proprotein convertase subtilisin/kexin type 9
PCE: Pooled Cohort Equations
PVD: peripheral vascular disease
REC: Research Ethics Committee
SCORE: Systematic Coronary Risk Estimation

## Acknowledgments

This research was conducted using the UK Biobank resource. We thank the participants of the UK Biobank. The work was performed under UK biobank project number 9310.

## Figure Legends

Figure 1 Centiles of Lp(a), along with 95% confidence intervals, by sex and ethnicity in the whole cohort. Red denotes white ethnicity, green denotes black ethnicity, blue denotes South Asian, and purple denotes other or mixed ethnicity.

Figure 2 Association of Lp(a) with outcomes after adjusting for classical risk factors among participants without baseline CVD and not taking a statin.

Figure 3 Population attributable fractions (with 95% confidence intervals) of Lp(a) over 100nmol/L for each outcome of interest within the whole cohort (orange), the primary prevention cohort (yellow) and the high risk cohort (history of CVD or taking a statin; green).

**Supplementary Table 1.** Association of Lp(a) (nmol/L) with outcomes of interest.

| Cohort/Outcome | No incident event | Incident event | P-value for trend |
| --- | --- | --- | --- |
| **Cohort with no baseline CVD or statins (n=340,333)** |  |  |  |
| Composite CVD | n=334,208<br>19.0 (7.6, 70.1) | n=6125<br>23.0 (8.6, 94.0) | <0.001 |
| Fatal CVD | n=338,706<br>19.1 (7.6, 70.5) | n=1627<br>23.1 (8.4, 91.9) | <0.001 |
| CHD | n=331,403<br>19.0 (7.5, 69.7) | n=8930<br>25.5(9.0, 101.2) | <0.001 |
| Ischaemic stroke | n=338,448<br>19.1 (7.6, 70.5) | n=1885<br>19.3 (7.5, 78.1) | 0.34 |
| Heart failure | n=339,495<br>19.1 (7.6, 70.5) | n=838<br>21.3 (7.6, 84.7) | 0.27 |
| Peripheral vascular disease | n=338,958<br>19.1 (7.6, 70.5) | n=1375<br>23.2 (8.7, 89.1) | <0.001 |
| **Cohort with baseline CVD or prescribed statins (n=73,391)** |  |  |  |
| Composite CVD | n=69,451<br>23.3 (7.9, 101.9) | n=3940<br>30.1 (8.8, 127.9) | <0.001 |
| Fatal CVD | n=71,771<br>23.5 (7.9, 102.8) | n=1620<br>28.2 (8.6, 127.7) | <0.001 |
| CHD | n=65,672<br>23.0 (7.8, 100.2) | n=7719<br>30.3 (8.8, 128.2) | <0.001 |
| Ischaemic stroke | n=72,085<br>23.6 (7.9, 103.2) | n=1306<br>26.8 (8.4, 110.8) | 0.091 |
| Heart failure | n=72,344<br>23.6 (8.0, 103.3) | n=1047<br>24.7 (7.4, 112.0) | 0.90 |
| Peripheral vascular disease | n=72,050<br>23.5 (7.9, 102.4) | n=1341<br>37.6 (9.1, 153.5) | <0.001 |

Values are n, median Lp(a) and (Q1-Q3)
CHD: coronary heart disease; CVD: cardiovascular disease; Lp(a): Lipoprotein (a)

**Supplementary Table 2.**
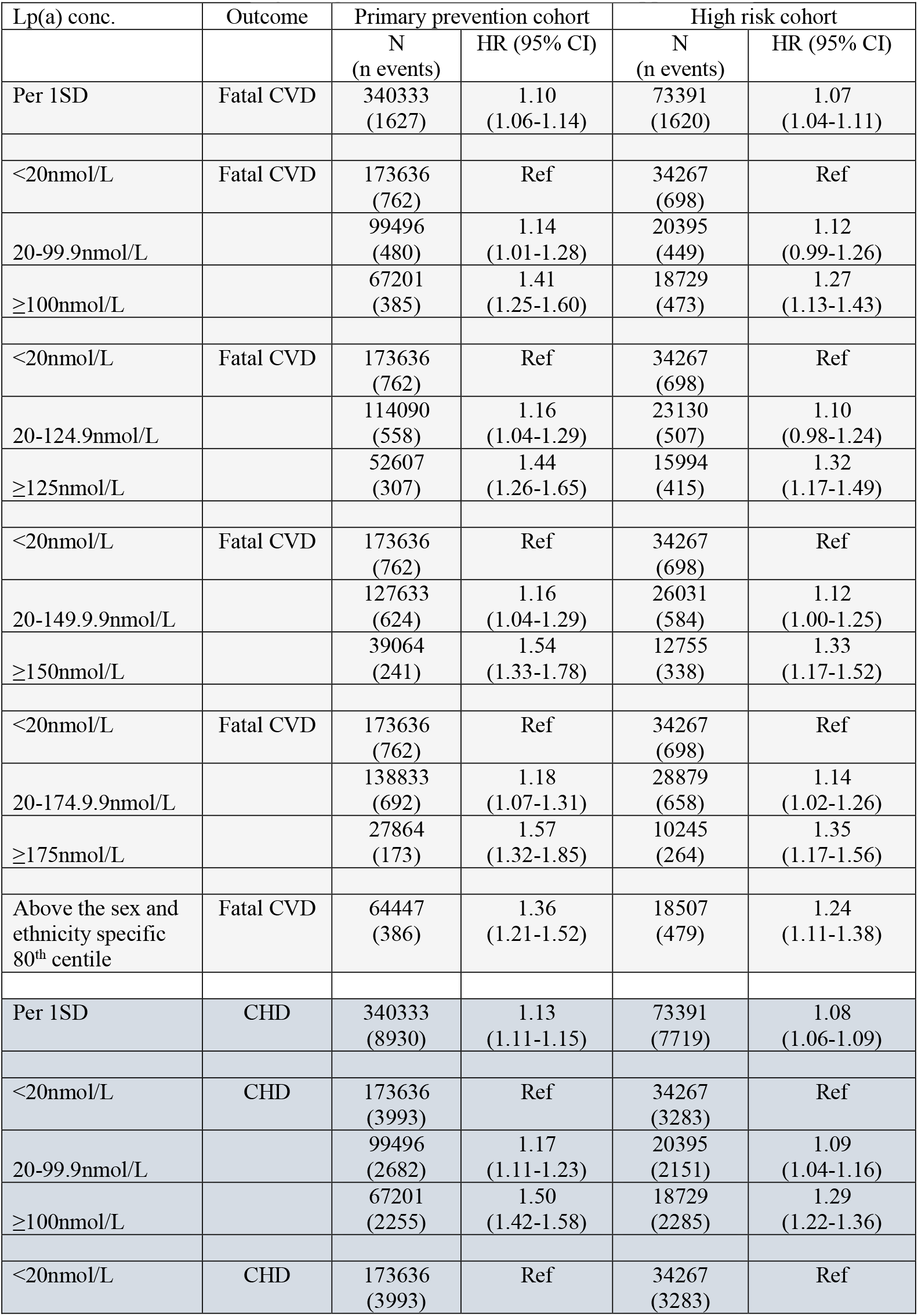
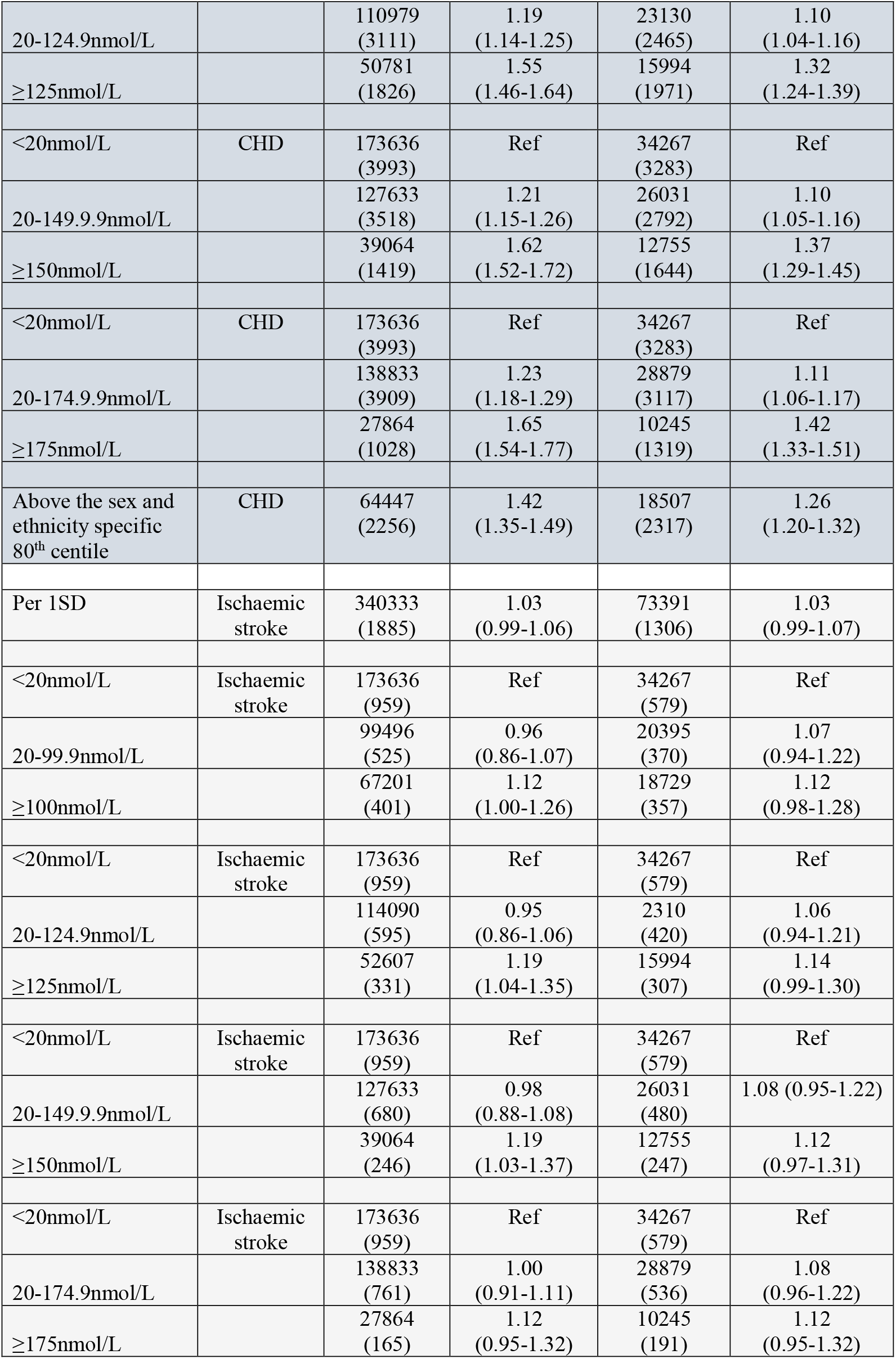
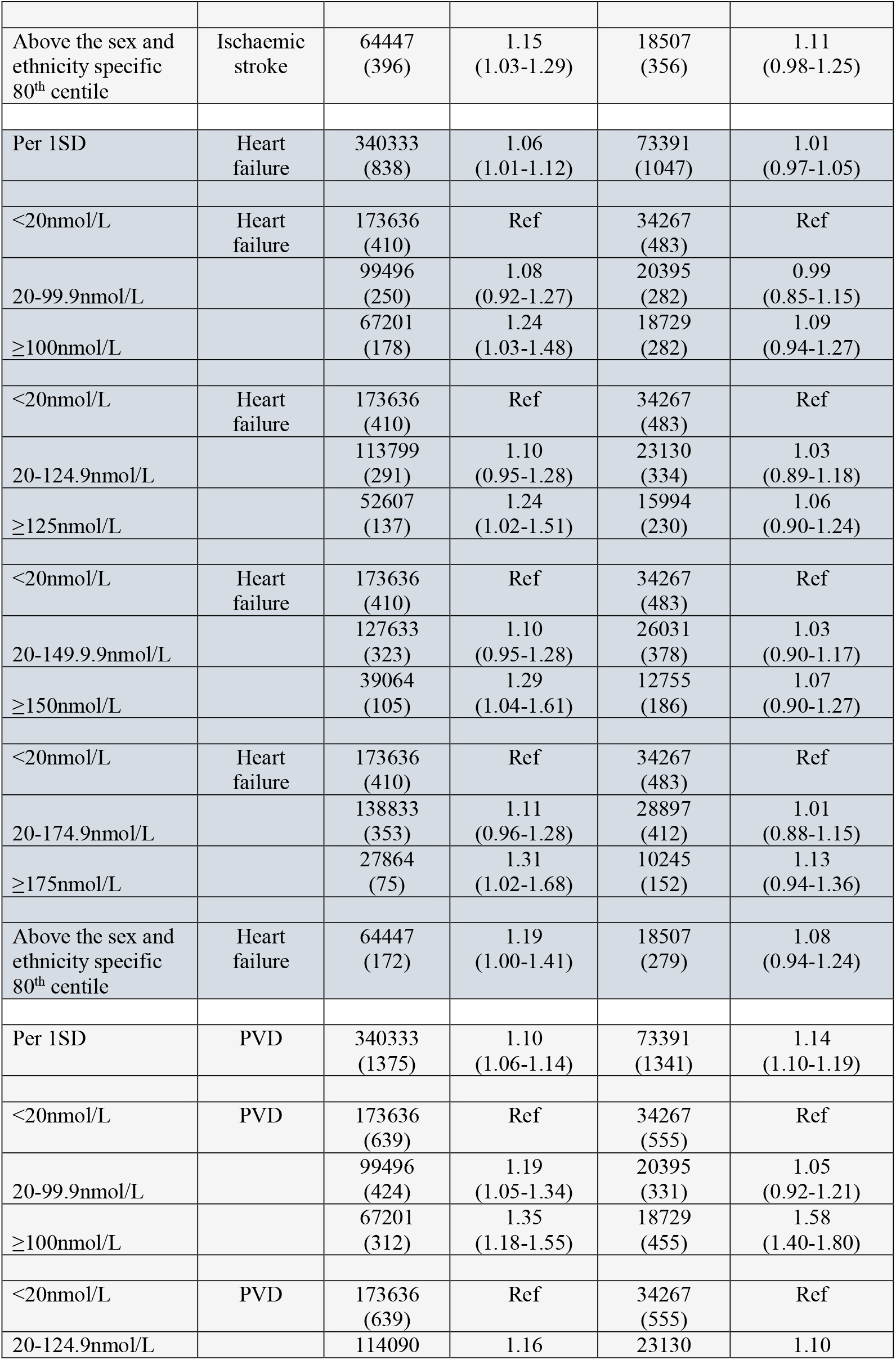
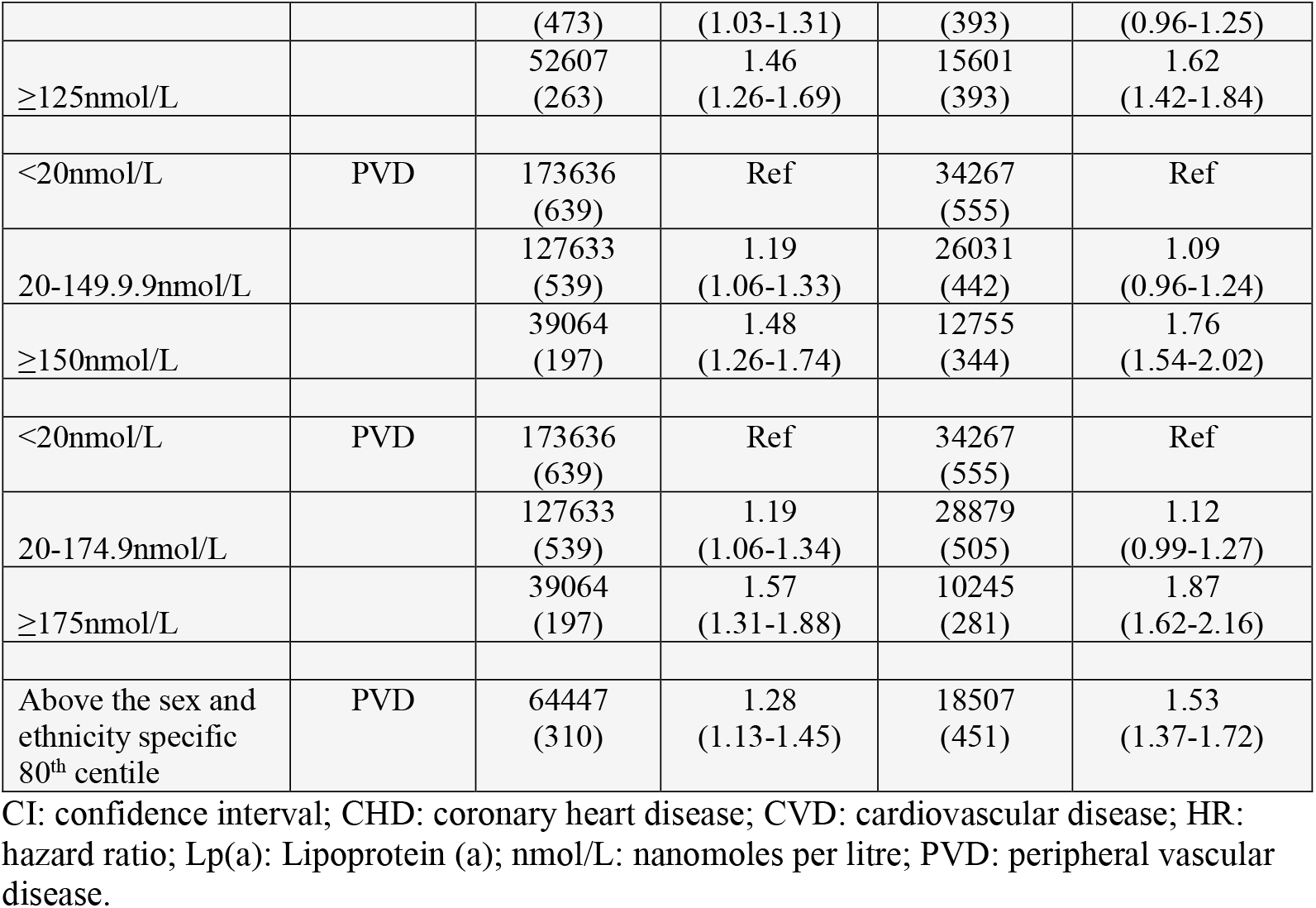
Association of Lp(a) (hazard ratio and 95% CI) as a continuous variable (per 1 SD increase in log Lp(a)) and as a categorical variable with secondary outcomes of interest after adjusting for classical risk factors in stepped models.

